# Feasibility of Non-Sedated Multispectral Neuroimaging in Newly Diagnosed Children with Leukemia

**DOI:** 10.64898/2026.04.21.26351310

**Authors:** Mary-Kaylin Tran, Lauren Appell, Simon Chung, Madison Sanders, Amy Jones, Mandy Fenwick, Elizabeth Oest, Xiawei Ou, Jason Farrar, Andrew W. Brown, Timothy R. Koscik, Ellen van der Plas

## Abstract

**Introduction:** Children with acute lymphoblastic leukemia (ALL) require neurotoxic treatment during critical periods of brain development. Multispectral magnetic resonance imaging (MRI) offers opportunities to characterize the impact of chemotherapy on the developing brain, but this has been constrained by concerns regarding feasibility of obtaining quality MRIs from these critically ill children at a young age. Our goal is to demonstrate a framework for improving the feasibility of non-sedated, multispectral MRI in newly diagnosed ALL patients.

**Methods:** Participants were enrolled from August 2023 through February 2026, and included newly diagnosed ALL patients and controls recruited from the community ages 3-10 years old. Participants completed a series of three non-sedated MRI without contrast on a 3T scanner over 12 months. Sequences included anatomical T1-weighted and QALAS sequences for quantitative mapping, diffusion weighted MRI (DWI), MR Spectroscopy (MRS), and resting-state functional MRI (rs-fMRI). A sequence was considered complete if data quality was sufficient for successful image processing. Mixed-effects logistic regression models were used determine if group and study visit were associated with MRI success rates across sequences.

**Results:** The sample included 17 ALL patients and 30 controls, who contributed to 38 and 75 observations, respectively. Of 113 attempted scans, 105 had ≥1 sequence completed (93%). Across assessments, ALL patients were less likely to complete a scan (31 of 38 attempts; raw rate: 82%) relative to controls (73 of 75; raw rate: 96%; model-estimated difference=-14.1%, 95%CI=-27.1%; -1.1%, p=0.033). Baseline success rate was 65% (11 of 17) for ALL patients and 93% (28 of 30) for controls (model-estimated difference=-26.8%, 95%CI=-52.7%; -0.9%, p=0.42); however, nearly all T1w scans in ALL patients were successful at follow-up visits (20 of 21; 95%), with no significant differences between groups (model-estimated difference at follow-up 1=-2.1%, 95%CI=-11.7%; 7.6%; model-estimated difference at follow-up 2=-8.3%, 95%CI=-26.3%; 9.6%). A similar pattern was observed for DWI, MRS and rs-fMRI sequences with relatively high completion rates at follow-up assessments.

**Discussion:** Non-sedated multispectral MRI is feasible in young children with newly diagnosed ALL, particularly with repeated visits and structured behavioral preparation. These results highlight opportunities to capture the scope and depth of neurodevelopmental changes concurrent with ALL treatment.

## Introduction

Therapy for acute lymphoblastic leukemia (ALL) is associated with neurocognitive impairment (1-3). Advanced, multispectral magnetic resonance imaging (MRI) methods can provide multi-faceted characterization of neurodevelopmental changes in critically ill children, provided that compliance challenges can be overcome (4-7). Given known risks (8-10), sedation is inappropriate in a research setting (11). Behavioral strategies enable non-sedated imaging in awake children (7, 12-14); however, clinical research frequently excludes the most medically complex cases due to perceived feasibility constraints (15-17). As a result, critically ill patients with ALL represent an overlooked population in the context of multispectral neuroimaging research. This lack of data creates a barrier to characterizing and mitigating neurotoxicity associated with high-intensity, life-saving interventions.

The current neuroimaging literature in ALL patients relies primarily on only two patient cohorts diagnosed 10 to 20 years ago (15, 16, 18-23), restricting its generalizability. Existing data also have limitations, as these studies included targeted neuroimaging strategies to identify leukoencephalopathy (18-21, 23). This narrow focus fails to capture the dynamic complexity of the developing brain, such as structural remodeling, functional network connectivity, and metabolic shifts. Leveraging the scope and depth of multispectral neuroimaging to track neurodevelopmental changes is key in advancing long-term outcomes in survivors. Additionally, by excluding children under age 7 and delaying assessments until the end of therapy, prior research has failed to capture the critical window of peak neurotoxic exposure and the developmental period of highest risk (15, 16). Closing these gaps requires the integration of multispectral neuroimaging during the early phases of treatment in young patients, a critical step for characterizing the immediate biological impact of therapy. The aim of the present study was to establish feasibility of applying non-sedated, multispectral, quantitative MRI to children with ALL (ages 3–10 years) through our on-going work characterizing the neurodevelopmental impact of ALL therapy. More broadly, this study establishes a framework for conducting high-fidelity neuroscience research in critically ill pediatric populations and evaluates MRI feasibility across patient groups and study visits.

## Methods

### Participants

Potentially eligible patients were identified at the Arkansas Children’s Hospital through electronic records between August 2023 through February 2026. Patients were eligible if they: 1) had T/B-cell ALL; 2) were between the ages of 3-10 years old at enrollment; 3) were treated with chemotherapy only.

An overview of how the sample was derived is shown in **Figure 1**. Patients were ineligible if they required planned cranial radiation and/or bone marrow transplantation; experienced relapse; had a major syndrome associated with intellectual disability (N=2); experienced severe head trauma; had MRI contraindications; or were unable to speak English (N=2). Of 33 ALL patients approached by February 2026, 21 consented to participate (64%). For those who did not participate, seven cited not having enough time, four indicated not being interested in participating in research, and one stated they did not think their child would tolerate the MRI. There were no significant differences in key demographic variables between individuals who consented and those who did not (**Table 1**).

**Figure 1.**
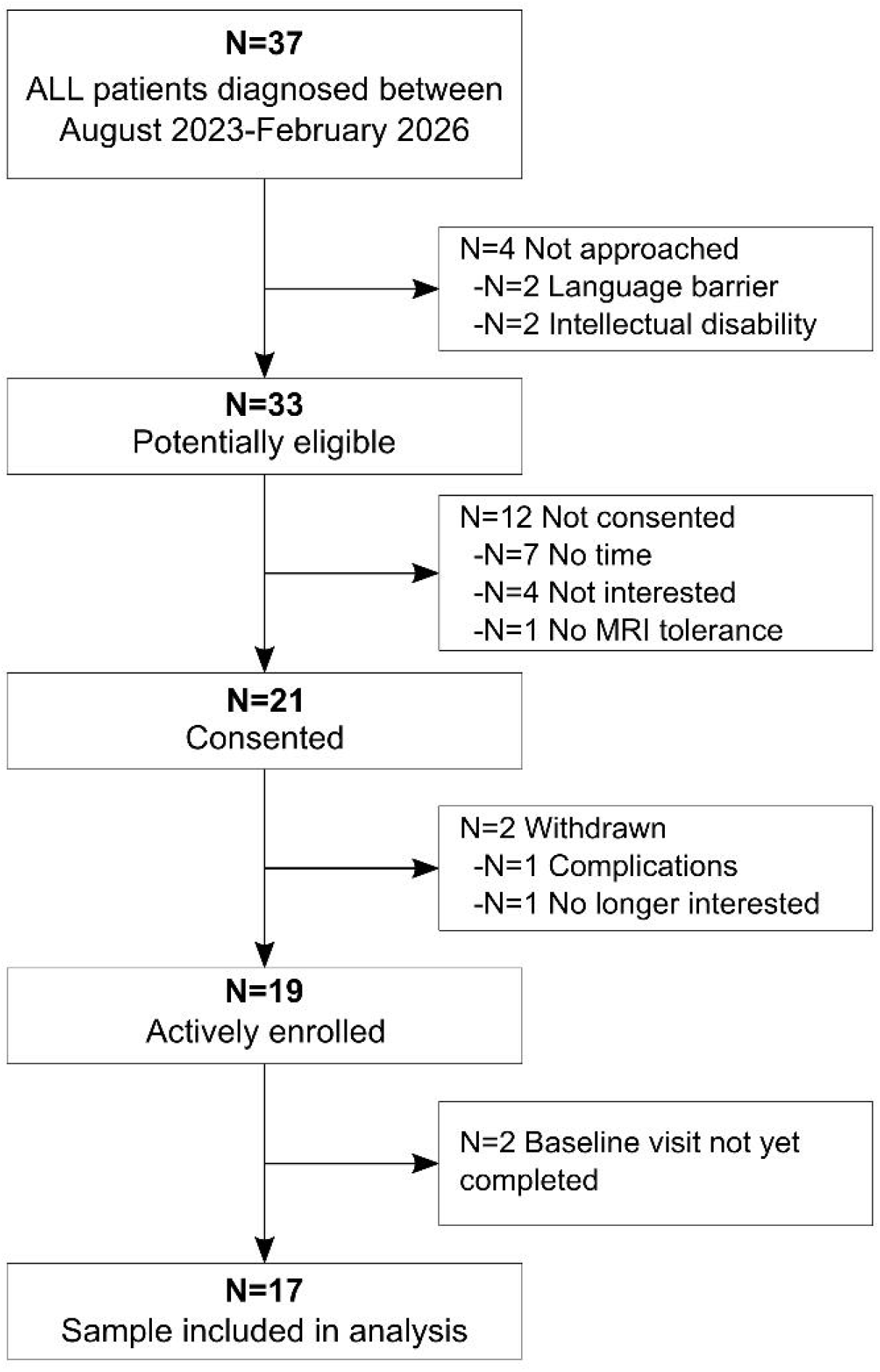
Diagram of patient selection.

**Table 1.**
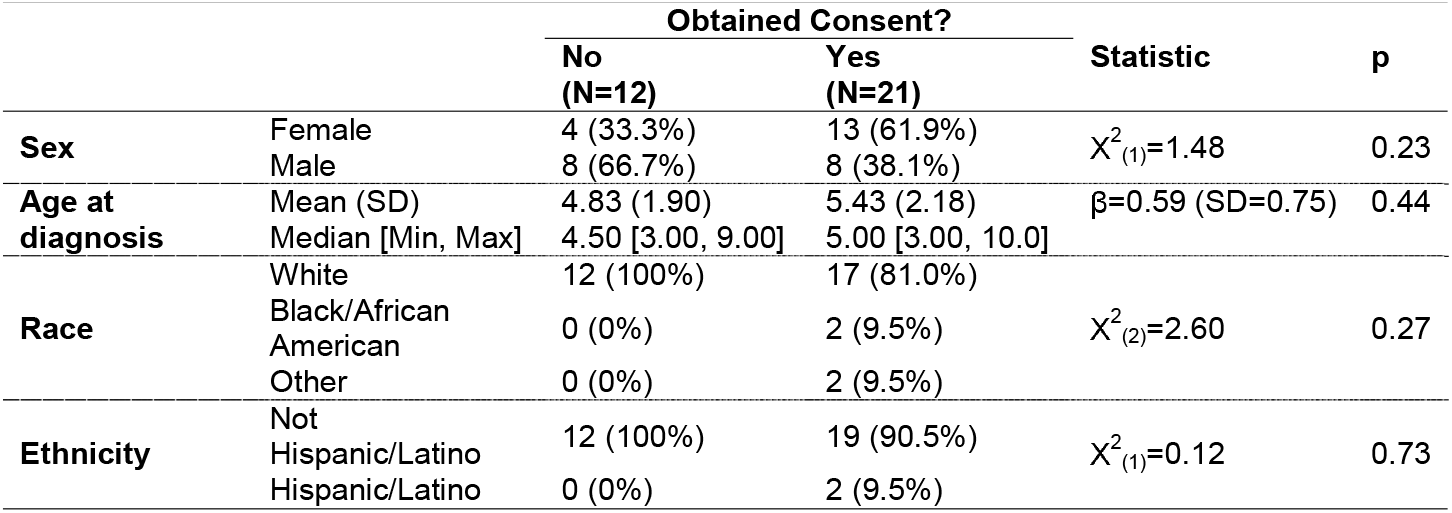
Demographics by consent status.

One patient was removed from the study due to complications that affected eligibility, and one patient withdrew from the study. Note that since this is an active study, not all enrolled participants have completed scheduled visits yet. This interim analysis validates the feasibility of our approach and establishes a methodological framework applicable to the study of diverse pediatric catastrophic illnesses.

Controls were recruited from the local community. Ineligibility criteria followed those of the ALL group, with additional exclusion of those in talented and gifted programs, to mitigate confounding neurocognitive differences.

Written informed consent was obtained from legal guardians, while participants provided either verbal (≥3 and ≤8 years old) or written assent (9-10 years old). The study was approved by the institutional review board of the University of Arkansas for Medical Sciences (IRB#275162).

### Study Procedures

As part of this ongoing study, participants complete three study visits over a one-year period **(Figure 2)**. At each study visit, participants completed neurocognitive testing (24-26) and neuroimaging. Parents completed surveys about family demographics (27), medical history, quality of life (28), and neurocognitive function (29). Concurrent with the study visit, blood samples were collected from participants to assess plasma markers of brain health, such as neurofilament light and glial fibrillary acidic protein (30-32). Residual CSF samples, obtained during standard-of-care lumbar punctures, are stored for subsequent proteomic and metabolomic profiling across the ALL treatment continuum (**Figure 2**).

**Figure 2.**
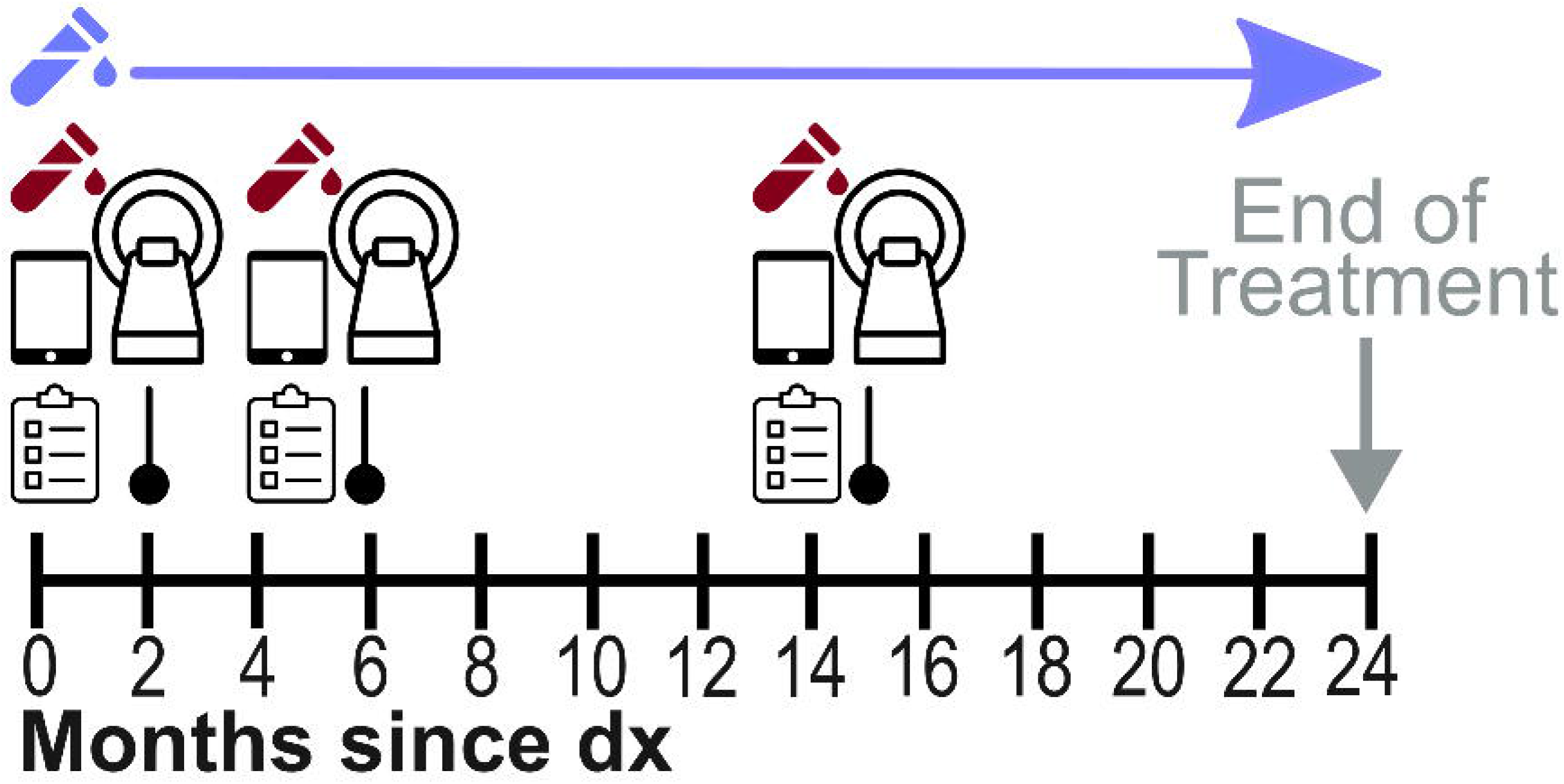
Study Procedures and data collection. Icons: cerebrospinal fluid (blue), blood (red), neurocognitive testing (iPads), parent surveys (clipboards), and MRI sessions. dx=diagnosis

Treatment for ALL follows a risk-stratified, phased protocol of varying intensity spanning more than two years (33). The most intensive therapy occurs in the months immediately following diagnosis; accordingly, study visits were aligned with key clinical milestones to ensure that patients followed comparable assessment timelines regardless of risk stratification (**Figure 3**). ALL patients completed the baseline assessment approximately 2 months post-diagnosis when medically stable. The second assessment occurred ∼4 months post-baseline, while the third assessment occurred 7-8 months following the second visit **(Figure 3)**. Study visits were typically scheduled around a clinic visit, with patients and families staying in a nearby hotel to minimize travel burden. MRI sessions were scheduled outside of blinatumomab cycles to avoid interruptions to the continuous steady-state infusion (34) and to bypass technical constraints associated with the MR-compatibility of the delivery equipment. Study visits were also scheduled outside of corticosteroid treatment windows to minimize the impact of medication-related behavioral dysregulation on compliance (35, 36).

**Figure 3.**
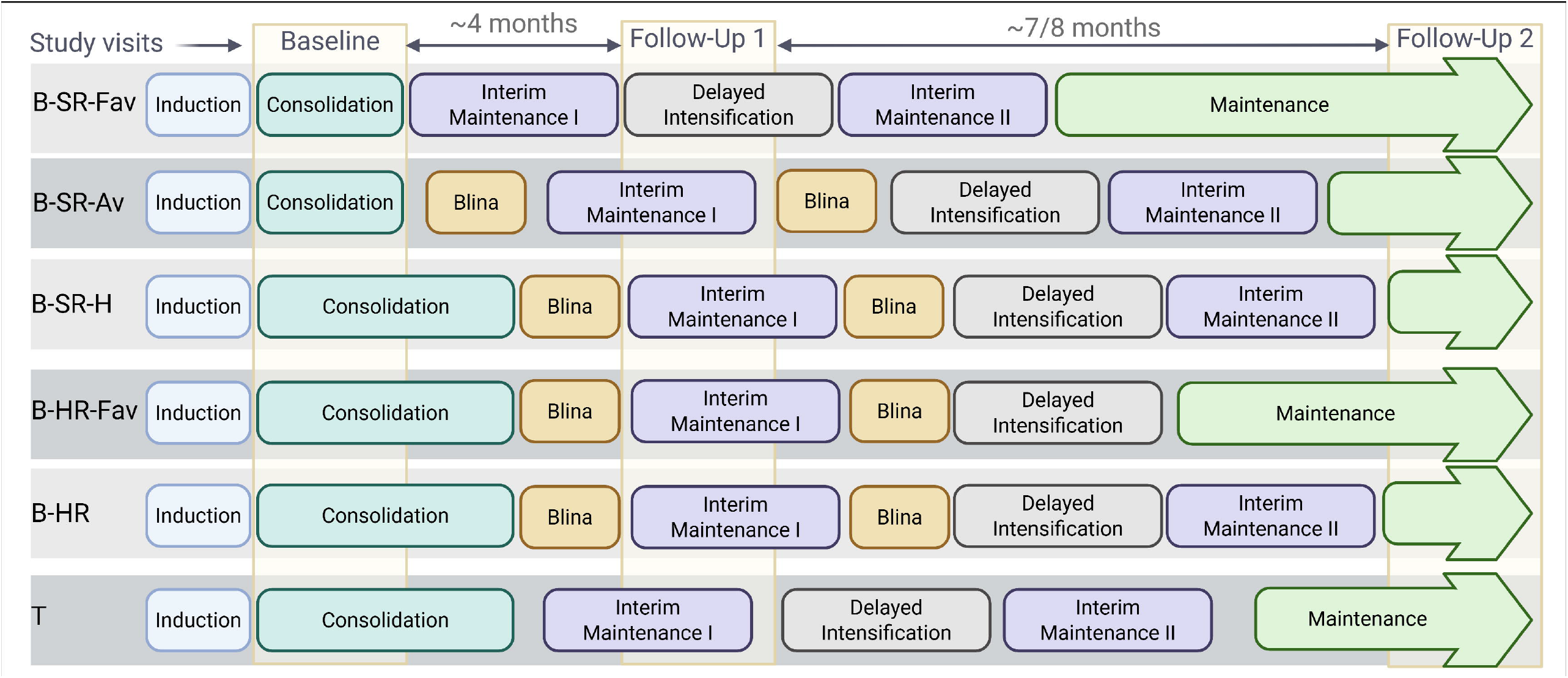
Study design and clinical treatment roadmap. The schematic illustrates the alignment of research assessments (Baseline, Follow-Up1; Follow-Up 2) with specific treatment blocks across B-ALL and T-ALL risk groups. Vertical shaded regions denote the assessment windows; colored rectangles represent discrete treatment phases. Abbreviations: B-SR-Fav, B-ALL Standard Risk Favorable; B-SR-Av, Standard Risk Average; B-SR-H, Standard Risk High; B-HR-Fav, High Risk Favorable; T, T-cell ALL. Created in BioRender.

### Neuroimaging

Participants completed a non-sedated MRI of the brain without contrast on a 3T Siemens Prisma scanner for research purposes only. Following recommended strategies for non-sedated scanning in young children (14), we used an experiential play based approach. Child life specialists or study team members used guided play with an MRI-themed Lego set or toy to familiarize children with scanning procedures prior to scanning. Participants practiced in a mock scanner (37), to familiarize with head coil placement and sounds. Participants could also use in-scan entertainment using an MRI-compatible audio-visual system, except during fMRI (see below). Noise-cancelling headphones were used to reduce noise.

Scan parameters were modeled after the national HBCD study (details found here: https://hbcdstudy.org/wp-content/uploads/2023/06/MRI-Parameters.pdf and (38)), and included (in order of acquisition): neuroanatomy (T1-weighted [T1w; 4:17 minutes [min] acquisition time], QALAS (38) for rapid, single scan whole-brain quantitative maps—T1, T2, and proton density; 4:14 min); white matter integrity and tractography (diffusion-weighted imaging, DWI; 2 x 6:24 min); neurometabolite concentration (magnetic resonance spectroscopy; 8:48 min); and resting-state functional connectivity (resting-state BOLD, rs-fMRI; 2 x 7:50 min). Self-chosen entertainment was discontinued at the start of the rs-fMRI sequence and replaced by a standardized computer-generated animation (39). Total scan time was approximately 45 minutes. As anatomical data are central to the study’s objective, the T1w sequence was designated as the highest priority, particularly when navigating behavioral challenges or limited participant compliance. Each sequence was considered complete if data quality was sufficient for successful processing through our image processing pipelines (**Figure 4**).

**Figure 4.**
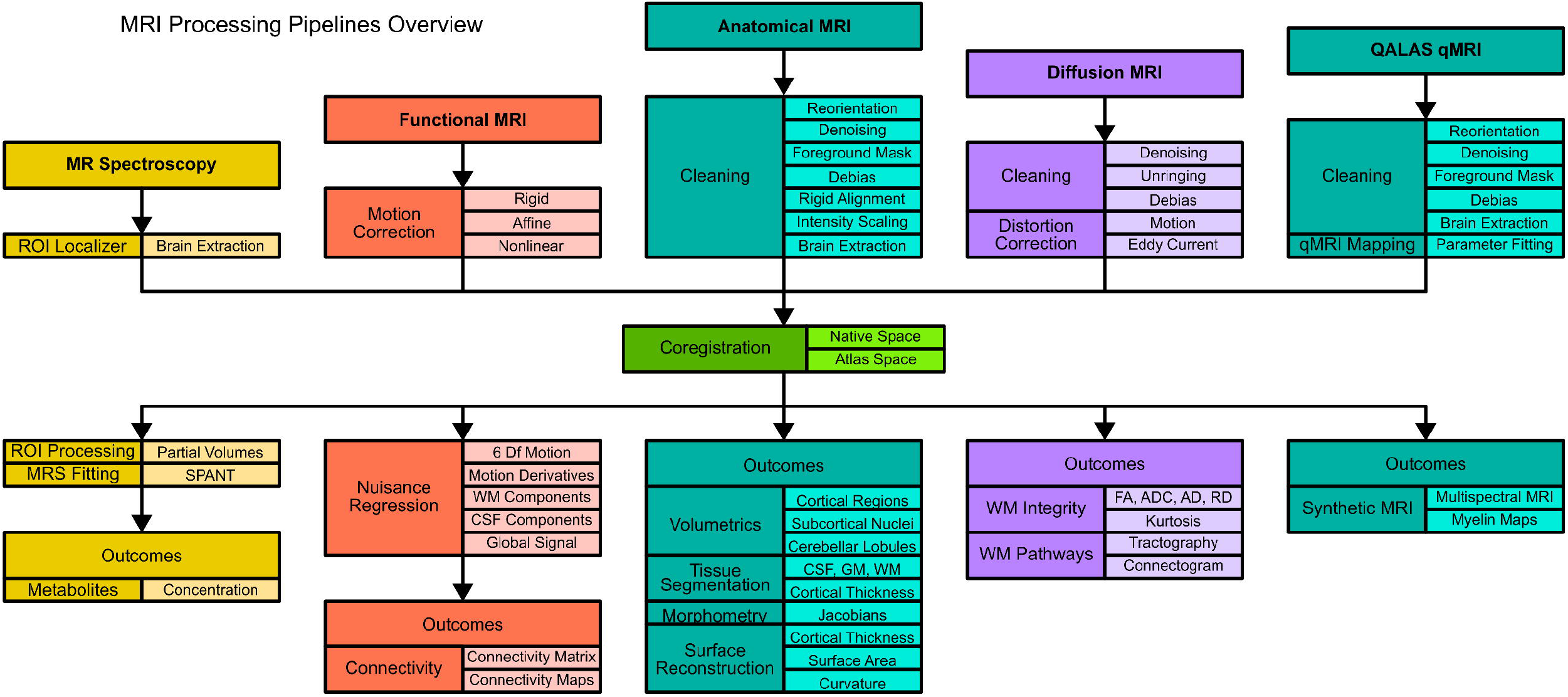
Summary of MRI processing pipelines.

### Statistical Approach

Group comparisons (Controls vs. ALL) on demographic variables were conducted with univariate regression or chi-square tests, where appropriate. For each MR sequence, we fit a mixed-effects logistic regression to determine if group and visit were associated with completion. Each model included a two-way interaction term between group and visit and random intercept for each participant. Standard maximum likelihood estimation could not be applied to the T1w and QALAS data, as the 100% compliance rate on the second attempt eliminated the statistical variability required for the model to converge. To stabilize the fixed-effects and random-effects estimate, we fit a Bayesian mixed-effects logistic regression with Cauchy distributions as the prior for fixed-effects (40), and Gamma distribution as the prior for the random effect covariance (41).

## Results

### Sample

The analytic sample included 30 controls and 17 ALL patients who completed 113 study visits (75 and 38 study visits in controls and patients, respectively). Baseline sample demographics are presented in **Table 2**. The sample was 59% female, with an average age at baseline of 6.96 years old (SD=1.99 years; range=3.21-10.94 years). No significant differences were observed in baseline demographics between groups. On average, patients underwent their first assessment 2.77 months post-diagnosis (SD=1.42 months).

**Table 2.**
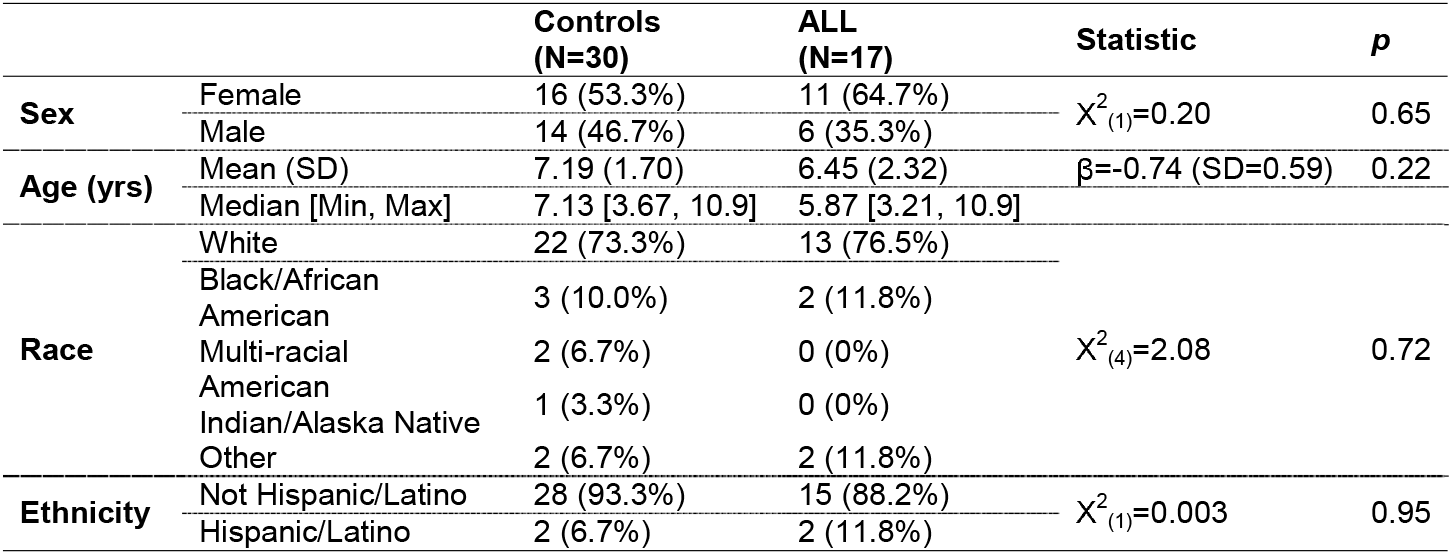
Baseline characteristics of the analyzed sample.

### Non-sedated MRI compliance

Of 113 attempted scans, 104 had ≥1 sequence completed (raw rate: 92%; **Figure 5**). Across study visits, ALL patients were less likely to complete scanning (raw rate: 82%) relative to controls (raw rate: 96%; model-estimated difference: T1w=-14.1%; 95%CI=-27.1%, -1.1%, p=0.003). Most missed sequences were due to participant refusal to complete the scan, with some exceptions noted below.

**Figure 5.**
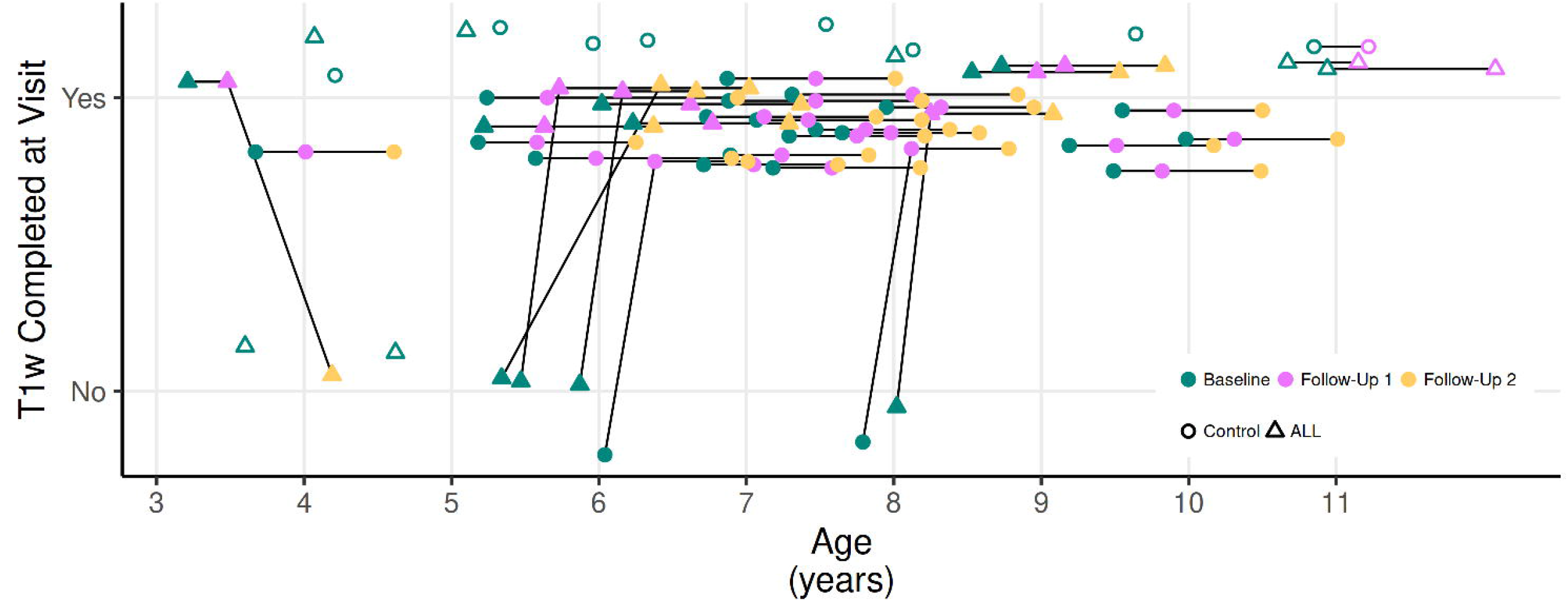
T1w completion for each participant. T1w success (y-axis) across age (x-axis) by visit (green=baseline; pink=follow-up 1; yellow=follow-up 2) and participant type (circles=controls; triangles=ALL patients). Open circles/triangles represent participant who have not completed the study yet.

Baseline assessments were least successful across sequences; however, compliance improved notably at subsequent study visits. Baseline T1w completion was 65% for ALL patients and 93% for controls, a difference that reached statistical significance (**Figure 6A; Table 3**). Of eight missed baseline scans (involving two controls and six ALL patients), seven participants cited anxiety about the procedure, and one ALL patient was unable to complete scanning due to dental caps. T1w scans at follow-up visits were almost always successful across both groups, with no statistical differences between patients and controls (**Figure 5; Figure 6A; Table 3**).

**Table 3.** Observed success rates across MR sequences, study visits and group.

| Sequence | Visit | Group | N | N<br>Success | % | Estimated<br>difference<sup>1</sup> | 95% CI of<br>Difference | p |
| --- | --- | --- | --- | --- | --- | --- | --- | --- |
| **T1w** | Baseline | ALL | 17 | 11 | 65% | -26.8% | 52.7%, -0.9% | 0.042 |
|  |  | Controls | 30 | 28 | 93% |  |  |  |
|  | Follow-up 1 | ALL | 11 | 11 | 100% | -2.1% | -11.7%, 7.6% | 0.672 |
|  |  | Controls | 23 | 23 | 100% |  |  |  |
|  | Follow-up 2 | ALL | 10 | 9 | 90% | -8.3% | -26.3%, 9.6% | 0.362 |
|  |  | Controls | 22 | 22 | 100% |  |  |  |
| **QALAS** | Baseline | ALL | 17 | 11 | 65% | -22.1% | -49.7%, 5.6% | 0.118 |
|  |  | Controls | 30 | 26 | 87% |  |  |  |
|  | Follow-up 1 | ALL | 11 | 9 | 82% | -16.0% | -39.8%, 7.7% | 0.186 |
|  |  | Controls | 23 | 23 | 100% |  |  |  |
|  | Follow-up 2 | ALL | 10 | 7 | 70% | -25.1% | -56.1%, 5.9% | 0.113 |
|  |  | Controls | 22 | 22 | 100% |  |  |  |
| **DWI** | Baseline | ALL | 17 | 11 | 65% | -22.3% | -51.7%, 7.0% | 0.136 |
|  |  | Controls | 30 | 26 | 87% |  |  |  |
|  | Follow-up 1 | ALL | 11 | 8 | 73% | -23.7% | -55.5%, 8.0% | 0.143 |
|  |  | Controls | 23 | 22 | 96% |  |  |  |
|  | Follow-up 2 | ALL | 10 | 7 | 70% | -21.2% | -55.7%, 13.2% | 0.227 |
|  |  | Controls | 22 | 21 | 96% |  |  |  |
| **MRS** | Baseline | ALL | 17 | 11 | 65% | -19.8% | -49.0%, 9.5% | 0.185 |
|  |  | Controls | 30 | 25 | 83% |  |  |  |
|  | Follow-up 1 | ALL | 11 | 8 | 73% | -23.1% | -53.6%, 7.3% | 0.137 |
|  |  | Controls | 23 | 22 | 96% |  |  |  |
|  | Follow-up 2 | ALL | 10 | 6 | 60% | -34.5% | -70.8%, 1.8% | 0.062 |
|  |  | Controls | 22 | 21 | 96% |  |  |  |
| **rs-fMRI** | Baseline | ALL | 17 | 7 | 41% | -38.5% | -80.2%, 3.3% | 0.071 |
|  |  | Controls | 30 | 21 | 70% |  |  |  |
|  | Follow-up 1 | ALL | 11 | 8 | 73% | -22.0% | -54.4%, 10.5% | 0.184 |
|  |  | Controls | 23 | 21 | 91% |  |  |  |
|  | Follow-up 2 | ALL | 10 | 7 | 70% | -18.4% | -51.4%, 14.6% | 0.274 |
|  |  | Controls | 22 | 20 | 91% |  |  |  |

<sup>1</sup>Estimated differences and confidence limits were derived from fitted models. P-values are raw, unadjusted p-values

**Figure 6.**
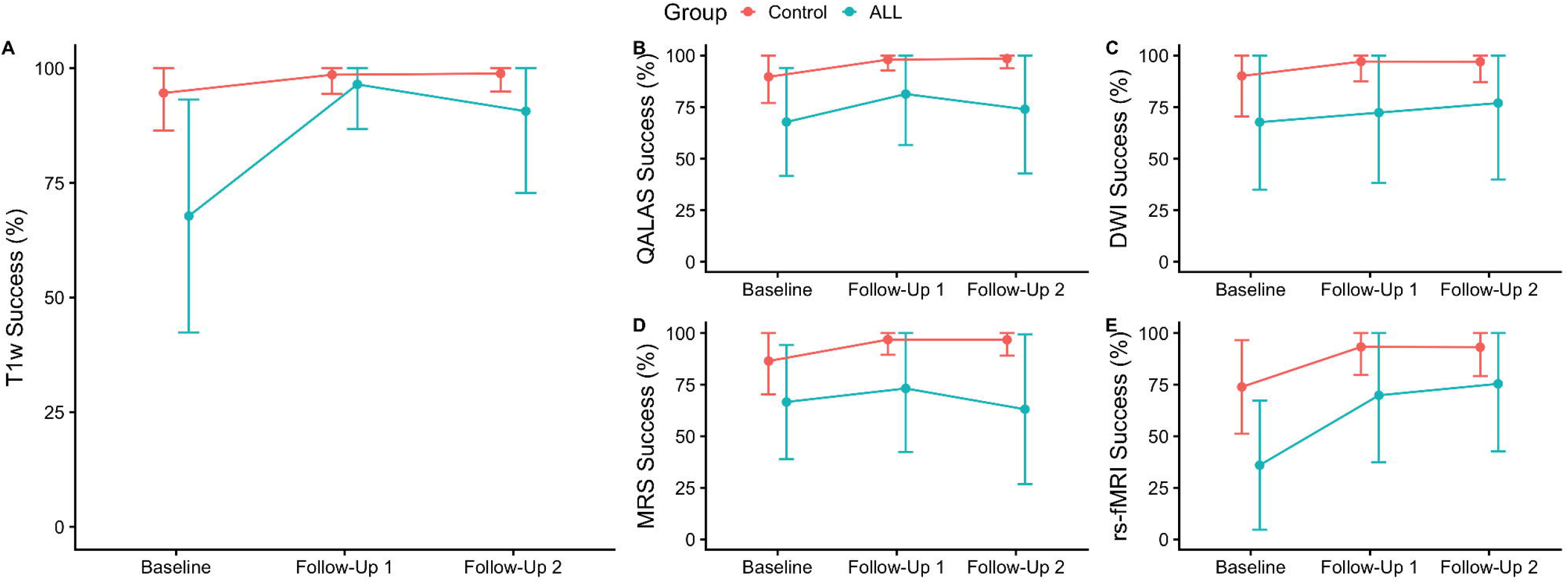
Model-derived scanning success rates across sequences. Estimated marginal percentages by group across study visits derived from the fitted model. Panels A-E display T1w, QALAS, DWI, MRS and rs-fMRI, respectively. Points represent estimated marginal means, and error bars indicate 95% confidence intervals.

A similar pattern where compliance improved with subsequent study visits was observed for the QALAS (**Figure 6B**), DWI (**Figure 6C**), MRS (**Figure 6D**), and rs-fMRI sequences (**Figure 6E**; **Table 3**). Success rates were lower in ALL patients than controls: the model-estimated difference between groups across study visits was 21.1% (95%CI=39.6%, 2.6%; p=0.03) for QALAS, 22.4% (95%CI=45.7%, 0.8%; p=0.06) for DWI, 25% (95%CI=45.9%, 4.0%; p=0.02) for MRS, and 27.8% (95%CI=54.1%, 1.6%; p=0.038) for rs-fRMI.

One QALAS sequence completed by an ALL patient at Follow-Up 1 was excluded for poor image quality. Technical difficulties resulted in the loss of three completed MRS datasets, including from one control at baseline, one control at Follow-Up 1, and one ALL patient at Follow-Up 2. The rs-fMRI appeared to be the most challenging sequence for participants to complete, with baseline success rates at 41% and 70% for ALL patients and controls, respectively (p=0.07; **Figure 6E**; **Table 3**). Equipment failure of the MR-compatible monitor precluded completion of two baseline assessments in ALL patients.

## Discussion

Although quantitative neuroimaging holds great promise for identifying markers of abnormal brain development in children with newly diagnosed ALL, such studies remain limited in part due to concerns about feasibility. This study demonstrates neuroimaging data can be collected from newly diagnosed young ALL patients, particularly with MRI acclimatization and individualized behavioral preparation. Prior studies focused on qualitative outcomes (18-21), or on older patients at the end of treatment (15, 16). We expanded these findings by demonstrating that various quantitative MRI methods can be applied early in treatment, and—with play-based methods, gradual acclimation, and repeated exposures—we can create opportunities to assess when and where neurodevelopmental changes occur. Combined with modern MR processing pipelines and evidence supporting the feasibility of administering cognitive assessments in ALL patients (42, 43), we have the tools to improve our understanding of the impact of neurotoxic treatments on the brain.

Compared to controls, ALL patients demonstrated lower completion rates, particularly at the initial neuroimaging session. Increased anxiety is well-documented in patients and survivors of childhood cancer (44, 45), which is consistent with our ALL patients who missed the baseline scan expressing anxiety as a predominant reason. The first study assessment occurred shortly after the initial ALL diagnosis and a more intense burden of medical interventions. Beyond psychological distress, these patients face unique clinical barriers, including intensive chemotherapy blocks that are incompatible with prolonged imaging, behavioral dysregulation triggered by neurotoxic agents, and the unpredictable nature of acute hospitalizations. Notably, our success rates for children with newly diagnosed ALL at baseline were slightly higher than recent reports of non-sedated neuroimaging in other pediatric populations, such as children aged 4–6 years with autism spectrum disorder or attention-deficit/hyperactivity disorder (65% vs. 54%) (46). Moreover, scan tolerance improved substantially by the second study visit. Our protocol intentionally avoided immediate re-scans following a failed session to allow the child to acclimate to the hospital environment. While this approach prioritized participant well-being, it resulted in missing data at the earliest developmental window. Despite incorporating play-based MRI acclimation that is known to increase compliance (47), the protocol restricted the study visit to a single day. Future studies might consider incorporating ‘a two-visit baseline model’, where an initial visit is dedicated to procedure familiarization followed by a separate session for data collection. However, the logistical challenges of scheduling additional visits for critically ill populations must be balanced against the potential for improved success rates.

To meet the study’s primary objectives, T1w sequences were prioritized, a strategy reflected in the comparatively lower compliance rates for subsequent acquisitions. The rs-fMRI sequence proved particularly challenging, likely due to its position at the end of the protocol and the necessity of pausing the participants’ visual entertainment. These varying success rates underscore the importance of optimizing scan duration and strategically prioritzing critical sequences early in the session. Indeed, a total scan time exceeding 45 minutes is a critical limiting factor to the success of non-sedated scanning in pediatric populations (7). Although non-sedated multispectral MRI presents logistical and behavioral challenges in pediatric critically ill populations, our findings demonstrate that high-quality data acquisition is achievable with structured preparation and repeated exposure.

Several limitations exist. First, as might be expected given the severity of the illness in our patient group, enrolling families in voluntary research is challenging. As a result, our sample size is relatively small, although the study is still actively enrolling participants. Notably, patients who did consent to participate did not differ substantially from patients who did not participate, partly mitigating risk of selection bias. Second, the technical complexity of multispectral MRI introduces vulnerability to data loss and transfer errors, underscoring the importance of standardized acquisition and data management procedures. Third, identifying an appropriate control group is difficult. We compared success rates against generally healthy controls. It is unclear if the differences in successful scanning would be similar between ALL patients and those with other conditions requiring intensive health interventions.

In sum, our findings highlight the practical and methodological advances that make early neuroimaging in young patients with ALL achievable. Despite challenges with enrollment and sequence completion, particularly for longer scans like DWI and rs-fMRI, our approach offers a viable framework for future studies. Careful session planning and continued emphasis on a developmentally appropriate play-based approach is essential to optimizing data collection. Integrating these imaging markers with neurocognitive data and biological samples provides a multimodal framework to elucidate the complex interactions between cancer, its treatment, and the developing brain. These insights lay important groundwork for longitudinal research aimed at capturing the evolving effects of treatment on the developing brain. More broadly, our work demonstrates the feasibility and utility of addressing clinically-relevant questions regarding neurocognitive late effects even within medically complex pediatric populations.

## Data Availability

All data produced in the present study are available upon reasonable request to the authors

## Acknowledgements

This work was funded by the National Cancer Institute (R37CA266135; PI E. van der Plas). Investigators were supported in part by the Arkansas Children’s Research Institute and the Arkansas Biosciences Institute. The views expressed in this article represent those of the authors and do not necessarily reflect the views of any organization.

